# Excess Mortality in the United States, 2020-21: County-level Estimates for Population Groups and Associations with Social Vulnerability

**DOI:** 10.1101/2024.01.14.24301290

**Authors:** Sasikiran Kandula, Katherine M. Keyes, Rami Yaari, Jeffrey Shaman

## Abstract

To assess the excess mortality burden of Covid-19 in the United States, we estimated sex, age and race stratified all-cause excess deaths in each county of the US during 2020 and 2021. Using spatial Bayesian models trained on all recorded deaths between 2003-2019, we estimated 463,187 (95% uncertainty interval (UI): 426,139 – 497,526) excess deaths during 2020, and 544,105 (95% UI: 492,202 – 592,959) excess deaths during 2021 nationally, with considerable geographical heterogeneity.

Excess mortality rate (EMR) nearly doubled for each 10-year increase in age and was consistently higher among men than women. EMR in the Black population was 1.5 times that of the White population nationally and as high as 3.8 times in some states. Among the 25-54 year population excess mortality was highest in the American Indian/Alaskan Native (AI/AN) population among the four racial groups studied, and in a few states was as high as 6 times that of the White population.

Strong association of EMR with county-level social vulnerability was estimated, including positive associations with prevalence of disability (standardized effect: 40.6 excess deaths per 100,000), older population (37.6), poverty (23.6), and unemployment (18.5), whereas population density (−50), higher education (−38.6), and income (−35.4) were protective.

Together, these estimates provide a more reliable and comprehensive understanding of the mortality burden of the pandemic in the US thus far. They suggest that Covid-19 amplified social and racial disparities. Short-term measures to protect more vulnerable groups in future Covid-19 waves and systemic corrective steps to address long-term societal inequities are necessary.

**Significance Statement:** All-cause excess mortality estimates, the difference between observed all-cause deaths and deaths expected in the absence of a pandemic, can help more fully assess the pandemic’s burden than direct Covid-19 mortality. Our estimates, based on a 17-year record of all deaths in the US and a Bayesian spatial model, quantify the differences in excess mortality across counties and by population age, race and sex, as well as between the first and second years of the pandemic. Furthermore, our results indicate that population-level socioeconomic indicators such as poverty, unemployment and educational attainment had considerable effect on excess mortality during the pandemic. Sustained efforts to protect vulnerable populations during future waves of Covid-19 (and other public health emergencies) remain vital.

## Introduction

Now in its third year, the Covid-19 pandemic has caused large-scale disruptions globally. As is often the case with systemic shock events, estimating the short- and long-term societal health effects of the Covid-19 pandemic is important yet difficult. One commonly used but incomplete measure in such cases is mortality, and in the context of Covid-19, following the standard used by most national surveillance systems, this translates to deaths with a confirmatory positive test for Covid-19. However, limiting mortality to Covid-19 certified deaths in the United States results in underestimates due to, in part, the limited testing capacity during the initial months of the pandemic. Additional grounds to expect underestimation of Covid-19 mortality exist. For example, periods of high pandemic severity have tested the capacity of death certification processes, potentially leading to inaccuracies in certification of cause of death. Similarly, due to the recognized inequities in access to testing and critical care (1, 2), the magnitude of the underestimation can differ by demographic group (race, age, sex, economic precarity), location and pandemic severity level (3, 4). These inequities are also likely to disproportionately affect mortality certification among vulnerable populations. Moreover, indirect effects of the pandemic, such as deaths due to limited access to preventive medical care and emergency procedures, increased drug overdoses, or domestic violence (5–8) would not be accounted when burden estimates are limited to certified Covid-19 deaths.

All-cause *excess* mortality estimates, defined here as the difference between observed all-cause deaths and deaths expected in the absence of a pandemic, can help quantify these effects and more fully assess pandemic burden. Excess mortality estimates have been previously reported in multiple contexts including assessment of the impacts of famines (9–11), wars (12), extreme weather events (13, 14) and climate change (15). Admittedly, excess death estimates do not capture intermediate effects of the pandemic that did not lead to death, such as effects of economic distress including homelessness (16), food insecurity (17), impact on child development (18), harm to mental health (19, 20), and the prolonged effects following acute infection (long-Covid) (21), and thus are an incomplete measure of the pandemic impact. Nevertheless, excess mortality estimates may provide a more comprehensive reckoning of deaths associated with the pandemic than cause-specific mortality estimates.

Excess mortality estimates during the pandemic have been reported for several countries. A recent comprehensive report from the World Health Organization (WHO) estimated 4.48 (95% credible interval: 3.93 – 5.07) million excess deaths globally in 2020 and 14.9 (13.3 – 16.6) million cumulative excess deaths by the end of 2021 (22, 23). Differences were also reported by WHO region with an estimated 1.25 (0.91 – 1.58) million excess deaths in Africa, 3.23 (3.16 – 3.30) million in the Americas, 1.08 (0.87 – 1.30) million in the Eastern-Mediterranean, 3.25 (3.18 – 3.32) million in Europe, 5.99 (4.50 – 7.72) million in South-East Asia and 0.12 (−0.65 −0.35) million in Western Pacific region through the end of 2021. Several other estimates for select countries, regions and for different pandemic waves have also been reported (24, 25).

In the United States, excess mortality estimates were reported as early as June 2020 (26, 27), and the US Centers for Disease Control and Prevention’s (CDC) National Center for Health Statistics (NCHS) provides frequently updated estimates nationally and at the state-level (28). Estimates for specific population subgroups (age, race, sex, ethnicity etc.)(4, 29–33) and geographically resolved estimates for states (34, 35) and select counties (36, 37) have also been reported, noting significant differences by demographic characteristics and geography.

This study adds to the existing literature on Covid-19 associated excess mortality in the US in three significant ways: a) while multiple national and state-level estimates have been reported, county-level estimates are relatively fewer and when available, estimates for a substantial proportion of the counties were missing or possibly unreliable due to the use of public versions of mortality datasets that suppressed counties with few deaths. Here, by making use of a nearly two decade-long historical individualized record of all deaths in the US, we were able to generate estimates for all counties in the US; b) we modeled 72 population subgroups separately (9 age, 4 racial and 2 sex groups), which enabled the capture of group-specific temporal trends and spatial patterns leading up to the pandemic, and hence, a better assessment of the differential impact of the pandemic on each subgroup; and c) the generation of county-level estimates allowed the analysis of the association between excess mortality and social inequities that predated the pandemic. SARS-CoV-2, like other pathogens, was superimposed on existing social and economic structures and hierarchies, influencing a person’s ability to limit exposure (by working-from-home, for example), transmission (by not residing in crowded or poorly ventilated housing), access testing when symptomatic, and access critical care when severely ill, all contributing to cascading downstream effects, in part measurable with excess deaths (38). Given the spatial heterogeneity of these combined effects, even within states, and the differential impact on demographic subgroups, excess mortality estimates presented here were hypothesized to be more reliable compared to models built with data at coarser granularity, and, consequently, supported more robust estimates of the effects of socioeconomic determinants.

In this study, we report the association of 18 socioeconomic indicators, individually and in composite, on county-level excess mortality during the first two years of the Covid-19 pandemic in the US. Assuredly, association analyses of excess mortality with *individual*-level socioeconomic attributes and *area*-level attributes both have their own unique value. In the absence of national level patient-level health records and/or registries, as is the case in the US, individual-level associations are harder to establish, but comprehensive measures of areal socioeconomic attributes in the US are readily available, and with necessary caution in interpreting associations from ecological analyses, they can serve as valuable counterparts to individual-level association studies.

To summarize, we used 72 strata-specific county-level models to estimate excess mortality in the US due to COVID-19 during 2020 and 2021, and demonstrate the bounds of new and emerging inequality among US counties and states across levels of social vulnerability, including:

`o` all-cause excess mortality in the US nationally, and excess mortality stratified by age, race and sex;
`o` all-cause excess mortality for the 50 states and the District of Columbia, overall and stratified by race and sex;
`o` all-cause excess mortality in 3142 US counties, stratified by race and sex;
`o` association between excess mortality and population social vulnerability at the county-level.

## Results

### National excess mortality estimates

We estimated median all-cause excess deaths of 463,187 (95% uncertainty interval (UI): 426,139 – 497,526) in 2020, and 544,105 (95% UI: 492,202 – 592,959) in 2021. These translate to 149 (95% UI: 137 – 160) excess deaths per 100,000 population in 2020 and 175 (95% UI: 159 - 191) excess deaths per 100,000 population in 2021. For comparison, 350,831 deaths during 2020 and 416,873 during 2021 were recorded to be from Covid-19 as the underlying cause. Based on the difference between certified Covid-19 deaths and estimated all-cause excess deaths, 112,356 (95% UI: 75,690 – 147,077) more deaths during 2020 and 127,232 (95% UI: 75,459 – 176,215) during 2021 occurred than would be expected from pre-pandemic trends and deaths directly attributed to Covid-19.

In 2020, excess mortality rates nearly doubled for each 10-year increase in age between the ages of 15 and 85 years, with the excess mortality rate for those older than 85 years nearly thrice that of 75-84 year olds (Figure 1). Excess mortality rates were higher in 2021 than in 2020 for all 10-year age groups between 25-84 years and lower for the 85+ age group (1969 per 100,000 in 2020; 886 in 2021). Excess mortality estimates were not significant in either year among those younger than 15 years.

**Figure 1.**
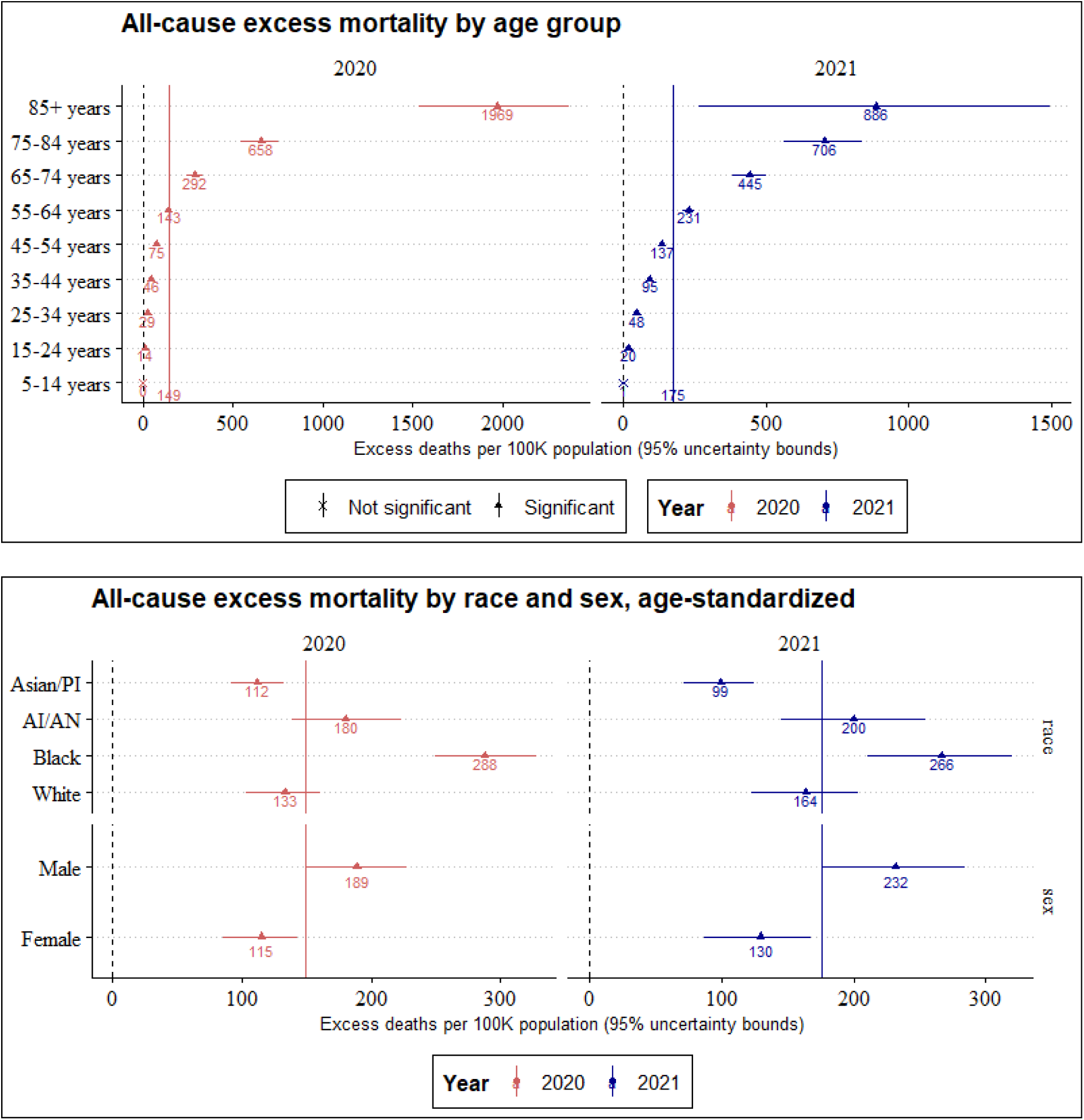
National excess mortality estimates in 2020 and 2021, stratified by: a) age; b) race and sex. Race and sex estimates are age-standardized to national age distribution using the direct method. The solid vertical lines denote national estimates. Instances where the lower 95% uncertainty bound was negative were not considered to be significant (denoted by a *x*; only occurred in 5-14 years).

Age-standardized excess mortality rates were higher in men than in women during both years, and this difference was larger in 2021 (189 per 100,000 among men and 115 among women during 2020; 232 and 130 respectively during 2021). The higher excess death rate among men was also consistent across all age groups (Figure ED1).

Age-standardized rates were estimated to be higher among the Black population relative to the White population during both years (288 among the Black population and 133 among the White population during 2020); this difference decreased in 2021 (266 and 164, respectively). Asian/Pacific Islanders had the lowest rates among the four racial groups modeled (112 in 2020; 99 in 2021) at approximately half the rate estimated for the overall population. Disaggregating excess rates by both age and race simultaneously showed considerable differences. While the overall rate among AI/AN was comparable to the White population, AI/AN had the highest excess mortality rate of the four race groups among the young and middle-aged (25-54 years) (Figure ED2). The rate differential of Asian/PI was also more pronounced in the young and middle-age populations (tending low) and was comparable to the White population in older age groups.

### Sub-national excess mortality estimates

During 2020, the highest excess mortality estimates were in states that experienced large outbreaks during the Spring wave (New York and New Jersey), and during late fall and winter wave of 2020 (Mississippi, North Dakota, South Dakota and New Mexico; Figure 2). Low or non-significant excesses were observed in the states of northern New England, the Pacific northwest, and in Alaska and Hawaii. Excess mortality rates were higher in 2021 than in 2020 in 36 of the 50 states and in DC (72.6%); the relative ranks of states were also considerably different (Spearman’s rank correlation: 0.27, *p* = .06). Large reductions were estimated in states in the Northeast (New York, New Jersey, Massachusetts, Connecticut) and large increases in excess mortality were estimated in Alaska, West Virginia, Arkansas, Kentucky and Tennessee.

**Figure 2.**
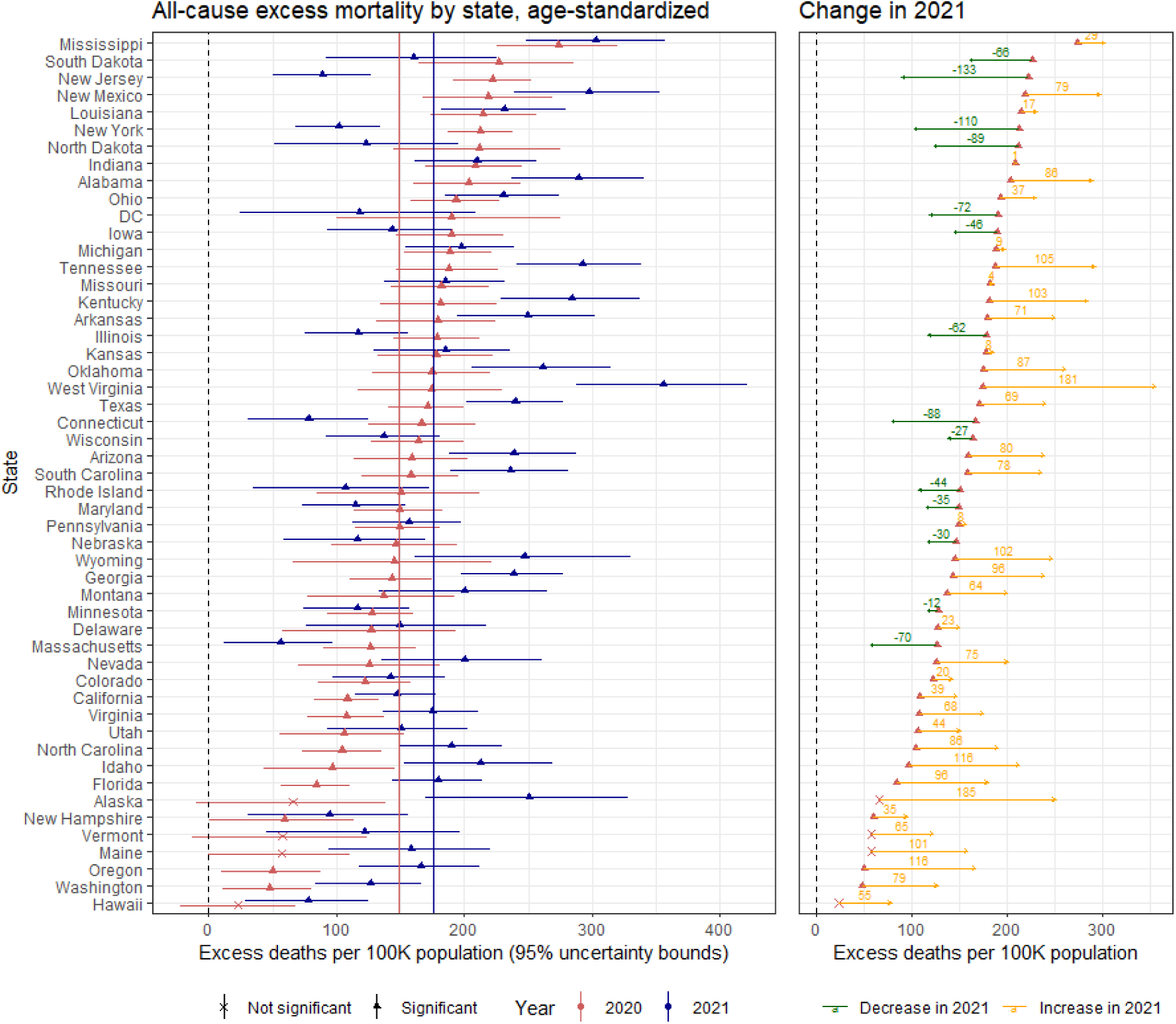
Age-standardized excess mortality estimates for US states and DC in 2020 and 2021. The states are ordered by median excess estimate for 2020, least to highest. The solid vertical lines denote national estimates for 2020 (red) and 2021 (blue). Instances where the lower 95% uncertainty bound was negative were not considered to be significant (denoted by a *x*; for example, Hawaii in 2020).

In all states, men were estimated to have consistently higher excess mortality rates than women, but the ratio of these rates varied by state. During both years, California had one of the highest ratios of excess mortality in men relative to women (2.5, Figure ED3), along with a few other states with large urban populations (New York, New Jersey, Illinois). The smallest ratios were estimated for states in the South Atlantic region (North Carolina, South Carolina, Arkansas) and the mid-West. Similarly, considerable heterogeneity was observed when state excess mortality rates were stratified by race (Figure ED4). Highest ratios of excess mortality among the Black population relative to the White population were estimated in Iowa (3.8 in 2021), Wisconsin (3.6 in 2021; 3.2 in 2020), Illinois (3.3 in 2021), Michigan (3.2 in 2020) and Nevada (3.2 in 2020); highest ratio of excess mortality among the AI/AN population relative to the White population were estimated in South Dakota (12.1 in 2021, 7.6 in 2020), Montana (7.5 in 2020), North Dakota (6 in 2021), Minnesota (5.9 in 2021) and Alaska (5.7 in 2021).

At the *county*-level excess deaths were significantly higher during 2021 than 2020 for 35.8% of counties, while a significant decrease was estimated for 17.5% of counties. For the remaining 46.7% of counties, estimates were not found to be significant in one or both years and hence the direction of change could not be determined. Two sets of counties exhibited particularly elevated rates during 2021 – counties in southern West Virginia bordering northeastern Kentucky and Tennessee; and most counties in New Mexico, Arizona, southwestern Nevada and southern Oregon (Figure 3).

**Figure 3.**
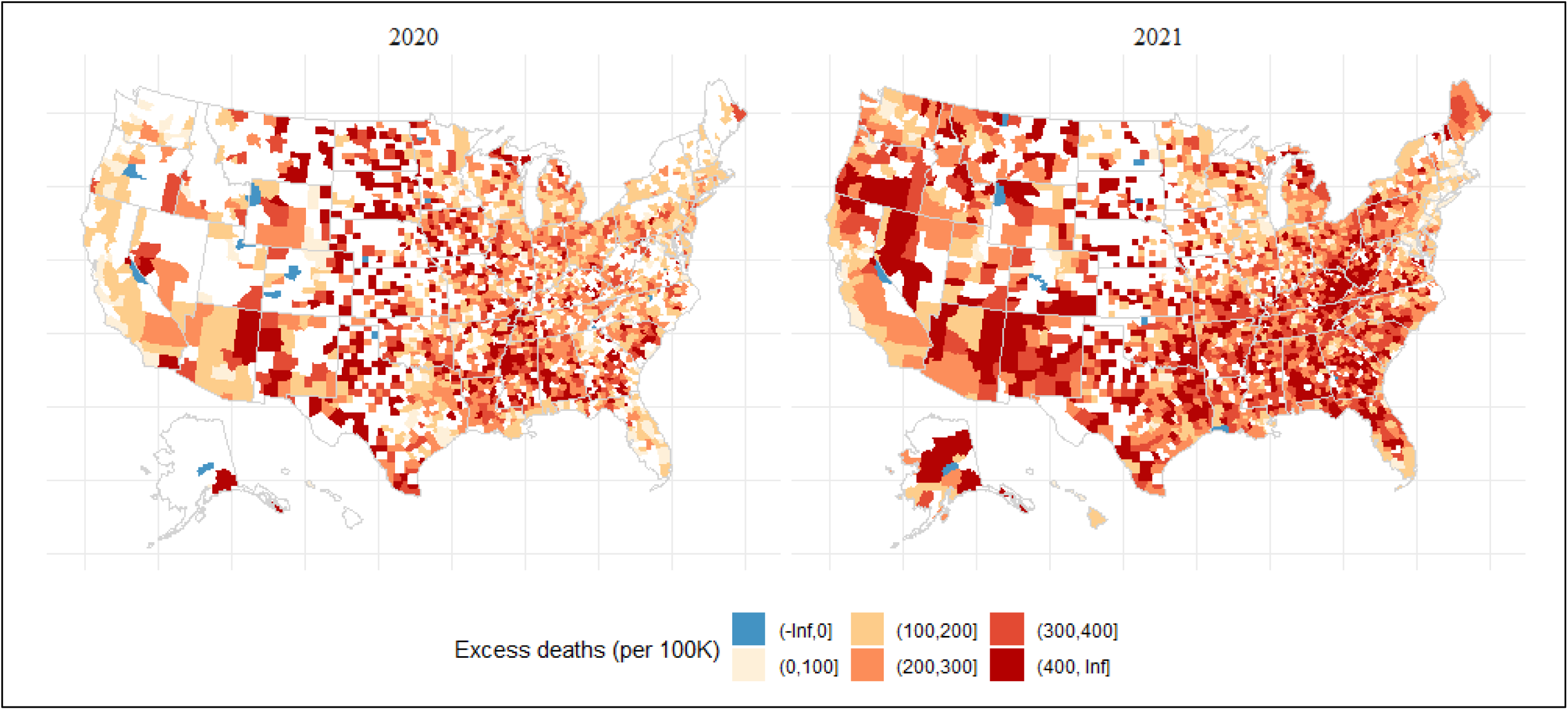
Map of excess mortality estimates for US counties in 2020 and 2021. Counties where the estimate was not found to be significant were suppressed (in white).

Disaggregating county-level excess mortality estimates during 2020 by sex of the decedent showed comparable spatial patterns between men and women, with particularly elevated rates in a majority of counties in Louisiana, Mississippi, Alabama, Texas, New Mexico and Arizona (Figure 4). A third of counties had statistically significant estimates in both sex groups, and in 65.5% of these counties men were estimated to have higher excess mortality than women. Disaggregating county-level excess mortality estimates during 2020 by race of the decedent showed that most excess deaths among the Black population were concentrated in a small number of states in the deep South. Similarly, high excess rates among AI/AN were concentrated in counties near Native American reservations (Oklahoma, Arizona, New Mexico). Disaggregating county-level estimates by race or sex for 2021 was not possible due to data suppression in the provisional data.

**Figure 4.**
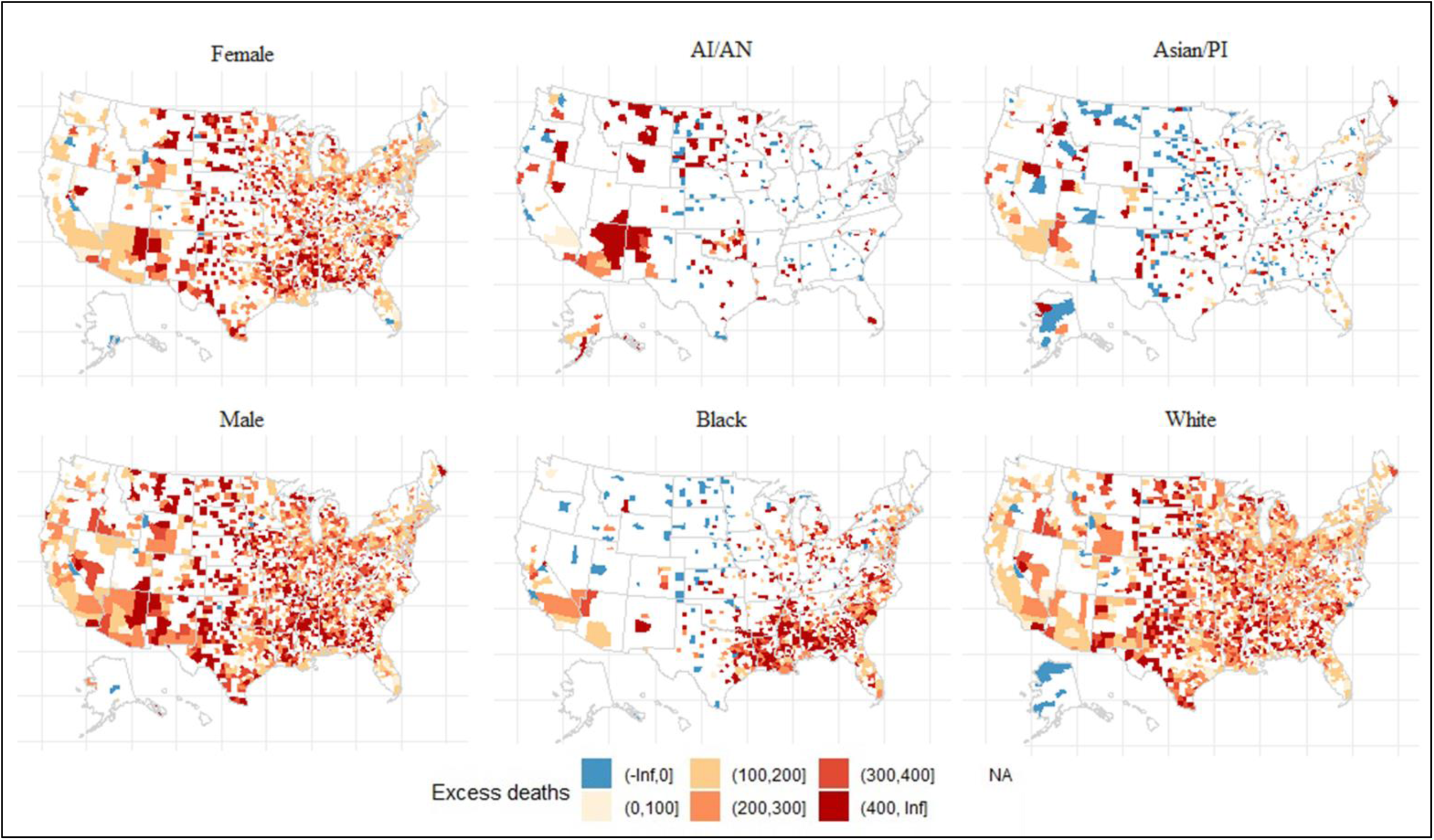
County-level excess mortality in 2020, stratified by sex (left column) and race. Counties where the estimate was not found to be significant were suppressed (in white).

### Associations between excess mortality and socioeconomic vulnerability

Linear univariate models, adjusting for deaths from Covid-19, showed strong associations of all-cause excess mortality rates with a majority of the social vulnerability measures considered. Limiting to measures with significant effect estimates in both years (*p* < 0.01), prevalence of individuals with disability (effect estimate per standard deviation: mean=40.6; 95% CI: 35-46), individuals over 65 years of age (37.6; 32-44), poverty (23.6; 18-29), mobile home residents (19.5; 14-25), unemployment (18.5; 13-24), and those who had not completed high school (15; 9-21) were estimated to have positive associations during 2020 (Figure 5). Population density (−50; −57 -−43), higher education (Bachelor’s degree or above; −38.6; −44 -−33), per capita income (−35.4; −41 -−30), and residence in buildings of 10 or more units (−28.9; −34 -−24) were found to be protective. Interestingly, prevalence of persons with limited English proficiency, proportion living in crowded accommodations, and proportion of minority population were also found to be protective, but with smaller effect sizes. Individually, these measures were found to explain between 49% and 54% of the variability in excess deaths (Figure 5).

**Figure 5.**
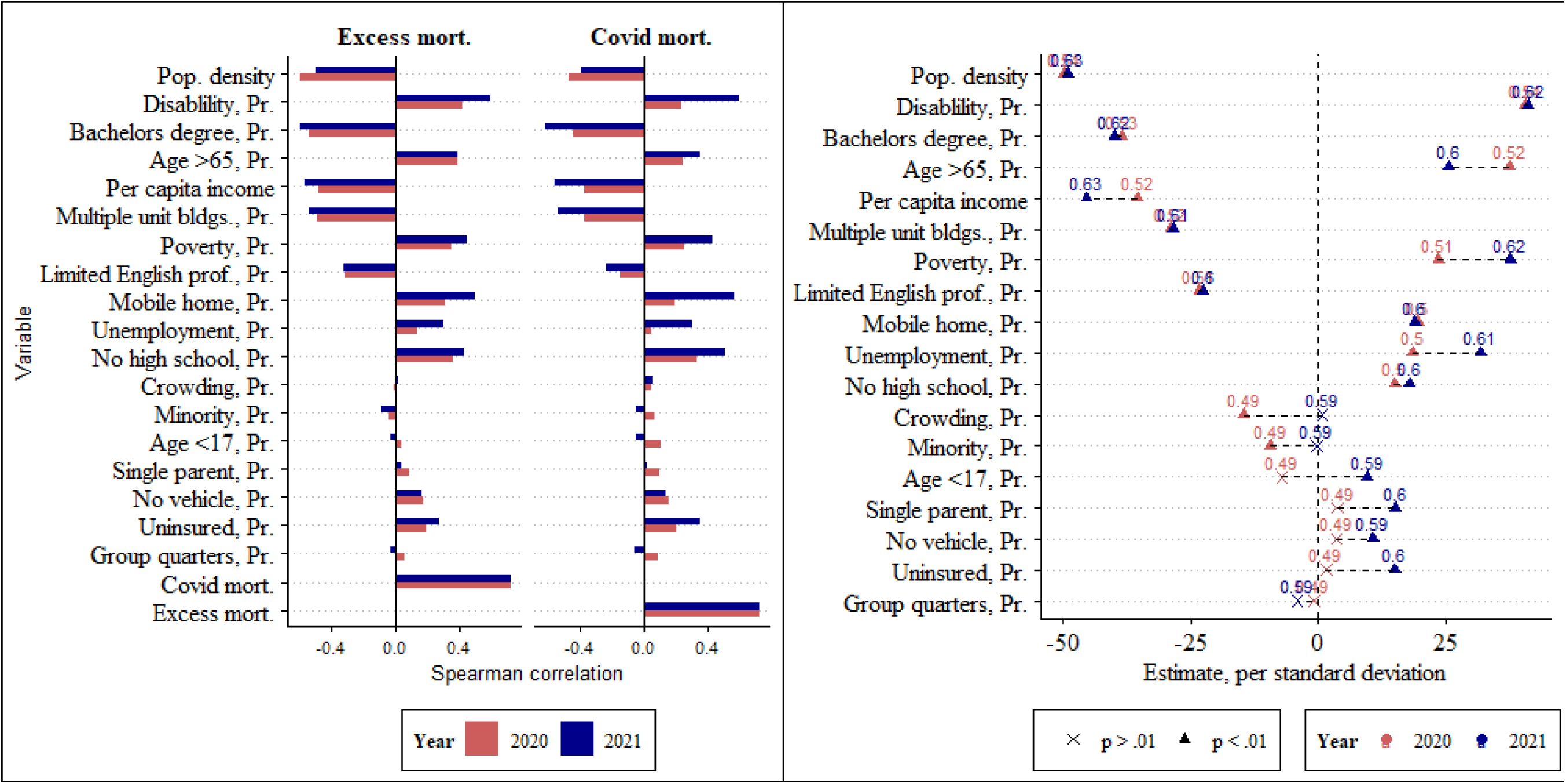
a) (left) Spearman correlation between social vulnerability measures and deaths from Covid−19 (*Covid mort.*) and all-cause excess mortality (*Excess mort.*) during 2020 and 2021; b) (right) Standardized effect estimates (per standard deviation) from linear univariate models of social vulnerability measures and excess mortality during 2020 (red) and 2021 (blue). Non-significant effect estimates are denoted by *x* (*p* > .01; for example, *Crowding Pr.* in 2021). The data points are labeled with estimated R^2^, and the measures are ordered by absolute effect in 2020, highest (*Pop. density, Pr.*) to least. See Table 1 for measure descriptions. Proportions are abbreviated as *Pr*.

The effect estimates of most measures were similar in 2020 and 2021, with four exceptions: increased positive associations of poverty (23.6 in 2020; 37.7 in 2021) and unemployment (18.5; 31.9), an increased protective effect of per capita income (−35.4; −45.5) and decreased positive association of prevalence of those over 65 years (37.6; 25.5). An increase in R^2^ for most measures was also observed (range: 59% to 63%).

Multivariate models that included all explanatory variables explained greater variability in both years (60% in 2020 and 67% in 2021, Figure 6). Only a third of the variables had significant effect estimates in each year, possibly due to the presence of considerable collinearities among the explanatory features (Figure ED5). Limiting to measures found to have significant effects in both years, a positive effect was estimated for unemployment (11.9 in 2020 and 17.6 in 2021), and a protective effect for population density (−42.5; −36.9), limited English proficiency (−19; −22) and prevalence of mobile home residents (−19.8; −16.8). A switch in the direction of association from the univariate model to the multivariate model in the latter measure is to be noted. Figure ED6 shows the multivariate model fit.

**Figure 6.**
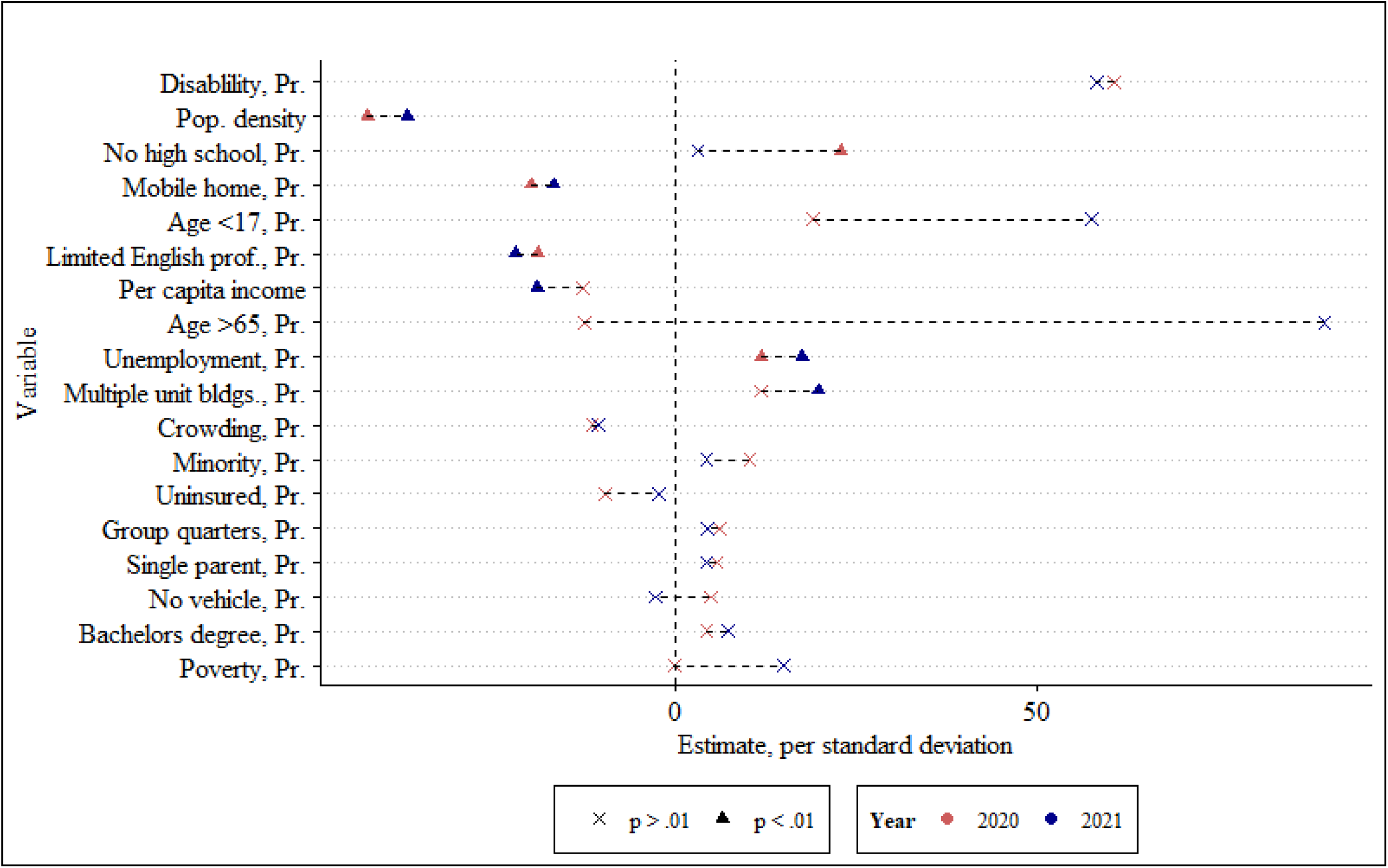
Standardized effect estimates (per standard deviation) from linear multivariate models of social vulnerability measures and excess mortality during 2020 and 2021. Non-significant effect estimates are denoted by a x (p > .01). Measures are ordered by absolute effect in 2020 highest (Disability, Pr.) to least, irrespective of statistical significance. See Table 1 for measure descriptions. Proportions are abbreviated as Pr..

**Table 1.** Measure of social vulnerability as available in CDC’s Social Vulnerability Index, 2018.

| ID | Variable label | Definition* | Median<br>(25%-75%) |
| --- | --- | --- | --- |
| 1 | Age >65, Pr. | Proportion of the population who are 65 years or older | 0.180 (0.15-0.21) |
| 2 | Age <17, pr. | Proportion of the population who are 17 years or younger | 0.223 (0.2-0.24) |
| 3 | Crowding, Pr. | Proportion of occupied housing units with more people than rooms | 0.019 (0.01-0.03) |
| 4 | Disability, Pr. | Proportion of civilian noninstitutionalized population who have a disability | 0.154 (0.13-0.18) |
| 5 | Group quarters Pr. | Proportion of population living in group quarters (nursing homes, correctional facilities, college dormitories etc.) | 0.020 (0.01-0.04) |
| 6 | Limited English prof., Pr. | Proportion of population (5 years or older) who speak English "less than well" | 0.007 (0-0.02) |
| 7 | Minority, Pr. | Proportion of the population who are minorities (all except White, non-Hispanic) | 0.161 (0.07-0.35) |
| 8 | Mobile home, Pr. | Proportion of housing that are mobile homes | 0.109 (0.05-0.19) |
| 9 | Multiple unit bldgs., Pr. | Proportion of housing that is in structures with 10 or more units | 0.029 (0.01-0.06) |
| 10 | No high school, Pr. | Proportion of the population (25+years) without high school diploma | 0.121 (0.09-0.17) |
| 11 | No vehicle, Pr. | Proportion of the population who do not have a vehicle | 0.056 (0.04-0.08) |
| 12 | Per capita income | Per capita income of the population, in 100,000 US dollars | 0.262 (0.23-0.3) |
| 13 | Pop. density | Population per sq. mile, log; 2010 census shape file | 1.651 (1.22-2.07) |
| 14 | Poverty, Pr. | Proportion of the population below poverty | 0.147 (0.11-0.19) |
| 15 | Bachelor's degree, Pr. | Proportion of population with Bachelor's degree or higher, 2016-20 ACS | 0.202 (0.16-0.27) |
| 16 | Single parent, Pr. | Proportion of households with single parent and children under 18 | 0.081 (0.07-0.1) |
| 17 | Unemployment, Pr. | Proportion of population who are unemployed | 0.054 (0.04-0.07) |
| 18 | Uninsured, Pr. | Proportion of total civilian noninstitutionalized population who are uninsured | 0.092 (0.06-0.13) |

\* data source is SVI based on 2014-18 ACS estimate, unless otherwise noted

Moran’s test (39) detected no autocorrelation in the residuals of the multivariate models, suggesting limited justification for using simultaneous autoregressive (SAR) models.

## Discussion

We have reported excess mortality estimates over 2 years at different spatial resolutions in the US, across multiple demographic groups, and analyzed the heterogeneity therein. Our estimates were based on a long historical dataset of all recorded deaths in the US, with no data suppression. The models employed were able to incorporate both temporal and spatial patterns specific to each demographic group. Together, we believe these provide a more reliable and comprehensive estimate of the mortality burden of the pandemic than previously reported, and add to the body of literature documenting the overall impact of Covid−19 in the US thus far.

We estimated a near doubling of excess mortality rate for each 10-year increase in age between 15 and 85 years, underscoring the heavy burden of the pandemic on the elderly. Excess mortality among the Black population was estimated to be 1.5 times that of the White population nationally, and as high as 3.8 times higher in some states. However, among the middle-aged (25−54 years) the American Indian/Alaskan Native (AI/AN) population had the highest excess mortality rates of the four race groups modeled, and in some states over 6 times that of the White population.

The differences in excess mortality among states were likely due to a complex interplay of the timing of introduction of the virus, regional characteristics aiding Covid−19 transmission (favorable humidity levels, high population density, type and quality of housing stock), ability of the disparate health systems to plan and respond to surges, the choice and timing of NPIs deployed in each state and population adherence to such measures, vaccination rollout and uptake, differences in underlying comorbidities, and the effects of concurrent public health emergencies.

A higher excess mortality was estimated for 2021 than 2020 in spite of the availability of vaccines, higher infection-acquired immunity, improved testing and treatment, and better know-how on the efficacy of different intervention measures. As the pandemic emerged during March 2020, reported estimates for 2020 were effectively for 10 months rather than a full year as in 2021 which might in part explain the lower rates. However a larger contribution to the increase in 2021 relative to 2020, is likely to have been from causes whose demand-side drivers were exacerbated by the pandemic, such as drug overdoses, alcohol-induced deaths etc. (40, 41). The approach described here could be used to generate cause-specific excess mortality estimates by training the models with cause-specific mortality during the pre-pandemic period as opposed to all-cause deaths. Such estimates would quantify the contributions of specific causes to excess mortality over the course of the pandemic, and the heterogeneity across regions and demographic groups.

The identified protective effect of higher per capita income and higher education were expected and may represent employment that more easily transition to remote work. Similarly, increased risk from unemployment, poverty, disability and age were expected. The decrease in effect size of older population (age > 65 years) during 2021 could be due to the availability of vaccines and improved treatment options as well as risk avoidance by this age group. Population density and higher prevalence of multi-unit residential buildings which are both potentially indicative of urbanity were also found to be protective. The protective effects of indicators of minority status, limited English proficiency and crowded housing were unexpected, and we hypothesize that some of this association may be attributed to the younger age profile of the Central and South American immigrant community in the US and healthy immigrant effect (42, 43). Additional analyses on the interaction of these measures with age would be necessary to understand this association.

While there is no ground truth to evaluate the accuracy of the estimates from different approaches, we compared our excess mortality estimates with NCHS estimates to flag any anomalies (28). National NCHS’ estimates were found to be lower in both years compared to our estimates (133 vs 149 in 2020; 162 vs 175 in 2021, Appendix text 4). At the state level, 91% of the NCHS estimates were within the 95% uncertainty bounds estimated by our method and were well correlated (Spearman’s *rho*: 0.76 in 2020; 0.93 in 2021), but lower.

As an additional test of model performance we generated out-of-sample estimates for the 10 years between 2010 and 2019 (Appendix text 3). Our results suggest that the observed mortality in these years, at most aggregations, were in line with expected mortality. Estimates were biased higher among AI/AN population in the validation period, suggesting that the expected deaths in 2020 and 2021 for this group were also likely to be overestimates, resulting in an underestimated burden due to the pandemic in this group. Interpreting the excess mortality of AI/AN relative to other racial groups requires caution.

Limitations of the study include the use of provisional mortality data for 2021; use of 2020 population estimates for 2021 without correcting for the large mortalities observed in 2020 (44), particularly impacting the elderly (75+ years); simplistic racial groupings; greater model uncertainty in strata with small and sparse populations; and absence of indicators of comorbidities and other population health indicators, as well as measures of change in vulnerability due to the pandemic.

With a few alterations to the model formulation, the approach described here, can be used to generate weekly or monthly mortality estimates (24). Better temporally resolved mortality estimates may help assess the effectiveness of deployed control measures and inform public health actions. For estimates at such finer temporal resolutions, retaining the 72 population groups may lead to sparser training datasets and biased model estimates, so fewer age strata might need to be considered. Additionally, the lag in public availability of detailed mortality data remains a barrier to generating more timely excess mortality estimates. Current mechanisms for the release of provisional estimates suppress data at finer resolutions, due to very legitimate privacy and quality concerns, but nevertheless have a negative impact on the timeliness of excess mortality estimates.

While short-term measures to protect more vulnerable groups in future Covid−19 waves are necessary, the inequities and societal drivers that created and sustain these inequities are long-term. Unless systemic corrective steps are undertaken, future emergencies, whether related to public health or climate, will continue to impose heavier tolls on the most vulnerable.

## Data and Methods

### All-cause mortality records

Detailed records of all-cause deaths for years 2003−2020 were obtained from the US National Vital Statistics System, and decedent’s age at time of death, race, sex, and county of residence extracted (45). From these individual records, for each county, annual count of deaths in 72 strata were computed, where a strata was defined by a combination of age, race and sex of the decedent using 9 age groups (5−14 years, 15−24, …, 75−84 years, 85+ years), 4 racial groups (White, Black, American Indian/Alaskan Native (AI/AN) and Asian/Pacific Islander (Asian/PI)) and two sex groups (Male and Female). Annual county-level mortality rates, defined as deaths per 100,000 population, were estimated using county population of each of the above 72 strata from the Bridged-Race Intercensal (2003−2009), decennial census (2010) and Postcensal (2011−2020) datasets (46, 47).

As detailed mortality data were not available for 2021 at the time this analysis was undertaken, provisional county-level all-cause death counts and rates (not stratified) were retrieved from the Multiple Cause of Death public interface on CDC WONDER (48). The interface suppressed counties with fewer than 10 deaths in 2021 (n=29; 0.9%), and assumed county population in 2021 to be identical to population in 2020.

Deaths directly attributed to Covid−19 were identified using *International Classification of Diseases−10* (*49*) underlying cause of death code U07.1, and similarly aggregated by county and strata.

### Measures of social vulnerability

The Centers for Disease Control and Prevention’s Social Vulnerability Index (SVI) is a composite index of 15 measures of community social vulnerability, resolved to county and census tract level (50). The measures were derived from the 5-year American Community Survey (ACS), with the most recent 2018 SVI based on 2014−18 ACS. See Table 1 for a list of measures and definitions. As some of the measures are protective (for example, per capita income), association of Covid−19 with the SVI’s overall index is hard to interpret, and instead here, we analyzed the association with each of the 15 measures separately.

Drawing on previously reported findings of strong associations with Covid−19 mortality, three additional measures were also included: population density (51) (persons per square mile (52), log transformed for normality), proportion of population with a Bachelor’s degree or higher (53) (B15003 variable of 2016−2020 ACS), and proportion of population who are uninsured (54).

Figure ED5 reports pairwise Spearman correlation among these 18 measures.

### Bayesian models of expected deaths

We trained 72 models, one for each population strata, using county-level annual deaths for the population in the strata during the period 2003−2019. Let *y*_*tsk*_ denote the number of all-cause deaths during year *t* in county *s* among the population in the *k^th^* strata, where a strata is defined by the combination of age, race and sex of the decedent; *P*_*tsk*_ and *r*_*tsk*_ are the corresponding population and risk. Under the assumption of a Poisson distribution for *y*_*tsk*_, the risk is modeled as:

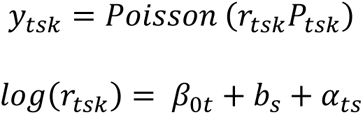

Here, *β*_0*t*_ is the time specific intercept defined as *β*_0*t*_ = *β*_0_ + *∈*_*t*_, with *β*_0_ representing the global intercept, and *∈*_*t*_, random effect representing the deviation of each year from *β*_0_, modeled as a first-order random walk process:

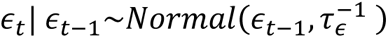

The spatial random effect *b*_*s*_ is modeled with an extension of the Besag-York-Mollie (55) model as

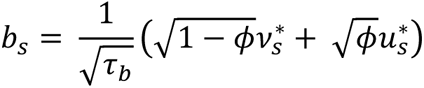

where 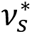 and 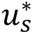 represent standardized (variance equal to 1) an unstructured random effect (*v_s_*) and spatially structured effect (*u*_*s*_), respectively; 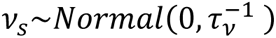 and

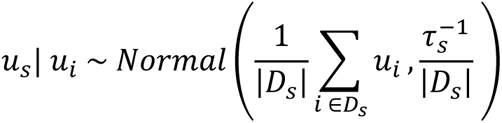

with *D*_*s*_ denoting the set of neighboring counties of county *s*, |*D*_*s*_| its cardinality, and *ϕ* a mixing parameter (0 ≤ *ϕ* ≤ 1) controlling the proportion of marginal variance explained by the structured effect. Finally, *α*_*ts*_ is a space-time interaction term modeled using independent and identically distributed Normal prior as 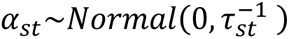.

Priors for hyperparameters *τ*_*∈*_, *τ*_*b*_, *τ*_*s*_, *ϕ* and *τ*_*st*_ were specified as penalizing complexity (PC) priors (56). To impose an upper bound on spatial relative risk based only on structured or unstructured variation at *exp*(2), the prior for 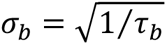 was specified as *Pr*(*σ*_*b*_ > 1) = 0.01 (24, 57). For the mixing parameter *ϕ*, as the relative contribution of the unstructured or structured effects to *b* is uncertain, a broad prior was specified with *Pr*(ϕ < 0.5) = 0.5. Priors for all other hyperparameters are identical to that of *τ*_*b*_. Prior for the fixed effect *β*_0_ was specified with a minimally informative Normal distribution.

### Posterior estimates and aggregation

To get estimates of expected deaths for each of 2020 and 2021 from each of the *k* models, Monte Carlo simulations were generated and 1000 random samples were drawn from the joint posterior distribution:

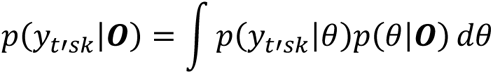

where *θ* is the (hyper) parameter vector as described above, and ***O*** the observed data through 2019.

The Integrated Nested Laplace Approximation (INLA) approach, as implemented in the *INLA* package (58, 59) in R (60), was used for Bayesian inference. To obtain posterior distributions of expected deaths for US nationally, or for a state or population subgroup and/or combinations thereof, a relevant subset of the *k* trained models was identified, and posterior distributions of location- and strata-specific expected deaths from this subset were aggregated. For example, to get county-level expected deaths among the White population, the posterior distributions of the 18 models (9 age and 2 sex groups) in which the strata was defined with *race = White* were summed. Similarly, to get estimates for a specific state, aggregation was limited to counties in the state.

Distributions of *excess* deaths were calculated as the difference between observed deaths and posterior distributions of expected deaths. Mean and 95% uncertainty bounds were computed from the posterior distribution of excess deaths. An estimate was considered to be significant if the lower (2.5%) and upper (97.5%) bounds had the same sign.

To adjust for differences in age profiles of the different population sub groups, the race and sex disaggregated excess mortality rates of US nationally, as well as the state-level rates, were age-standardized using the direct method (61, 62), with national age distribution as the reference population (see Appendix Text 2).

### Estimating association between excess mortality and social vulnerability

All-cause excess mortality can be seen as comprising of three components: a) deaths certified to be from Covid−19; b) deaths from Covid−19 infections that were not certified as such (due to limited availability of testing, stress on certification processes etc.); and c) excess deaths from all other causes, which can be interpreted as the indirect impact of the pandemic on mortality. We hypothesized that social vulnerability has had an effect on Covid−19 deaths but also on excess deaths not explicitly attributed to Covid−19 (i.e. the latter two components identified above), and that the magnitude of these two effects was different.

To estimate the association of select socioeconomic measures with all-cause excess mortality, we built linear univariate models with all-cause excess death rate as the response and each variable as the only explanatory variable, while adjusting for Covid−19 mortality rate. Separate models were trained for 2020 and 2021 and limited to counties with statistically significant excess deaths (62% of counties in 2020 and 74% in 2021). We report standardized effect estimates (per standard deviation) and the proportion of the variability explained by each variable in a year. As the SVI measures were estimated using 2014−18 ACS data, and are time-invariant, they reflect vulnerability at the start of the pandemic, and not the consequences of the pandemic.

Additionally, we built two linear multivariate models, for 2020 and 2021 respectively, with excess mortality rate as the response and all 18 socioeconomic measures together as explanatory variables, adjusting for Covid−19 mortality rate. We checked for the presence of spatial autocorrelation in these measures to inform whether approaches that explicitly model spatial autocorrelation, such as simultaneous autoregressive models (SAR) (63), were required.

We report effect estimates with Covid−19 mortality in Appendix Text 6.

## Data Availability

County-level mortality data for 2003−21, excess mortality estimates from this study for 2020 and 2021 and data dictionary are included as supporting information, and can be shared on public repository upon publication. County-level social vulnerability indicators are publicly available and can be downloaded from: https://www.atsdr.cdc.gov/placeandhealth/svi/data_documentation_download.html.

## Acknowledgements

This work is supported by a grant from the US National Institute of Mental Health (R01-MH121410), a contract from the US Centers for Disease Control and Prevention (75D30122C14289), and a gift from the Morris-Singer Foundation.

## Author Contributions

Designed research: Kandula, Keyes, Shaman

Performed research: Kandula, Keyes, Shaman

Analyzed data and developed tools: Kandula

Wrote the paper: (initial draft) Kandula; (revision) all authors

## Classification

Social Sciences (Psychological and Cognitive Sciences); Physical Sciences (Applied Mathematics); Biological Sciences (Medical Sciences)

