## supplemental text and figures for "Excess Mortality in the United States, 2020-21: County-level Estimates for Population Groups and Associations with Social Vulnerability"

**Classification:** Social Sciences (Psychological and Cognitive Sciences); Physical Sciences (Applied Mathematics); Biological Sciences (Medical Sciences)

**Keywords:** Covid-19; excess mortality; socioeconomic determinants; spatial models;

**This PDF includes:**

Extended figures (ED1-ED6)
Appendix texts (1-6)
Supplementary Figures (S1 to S5)

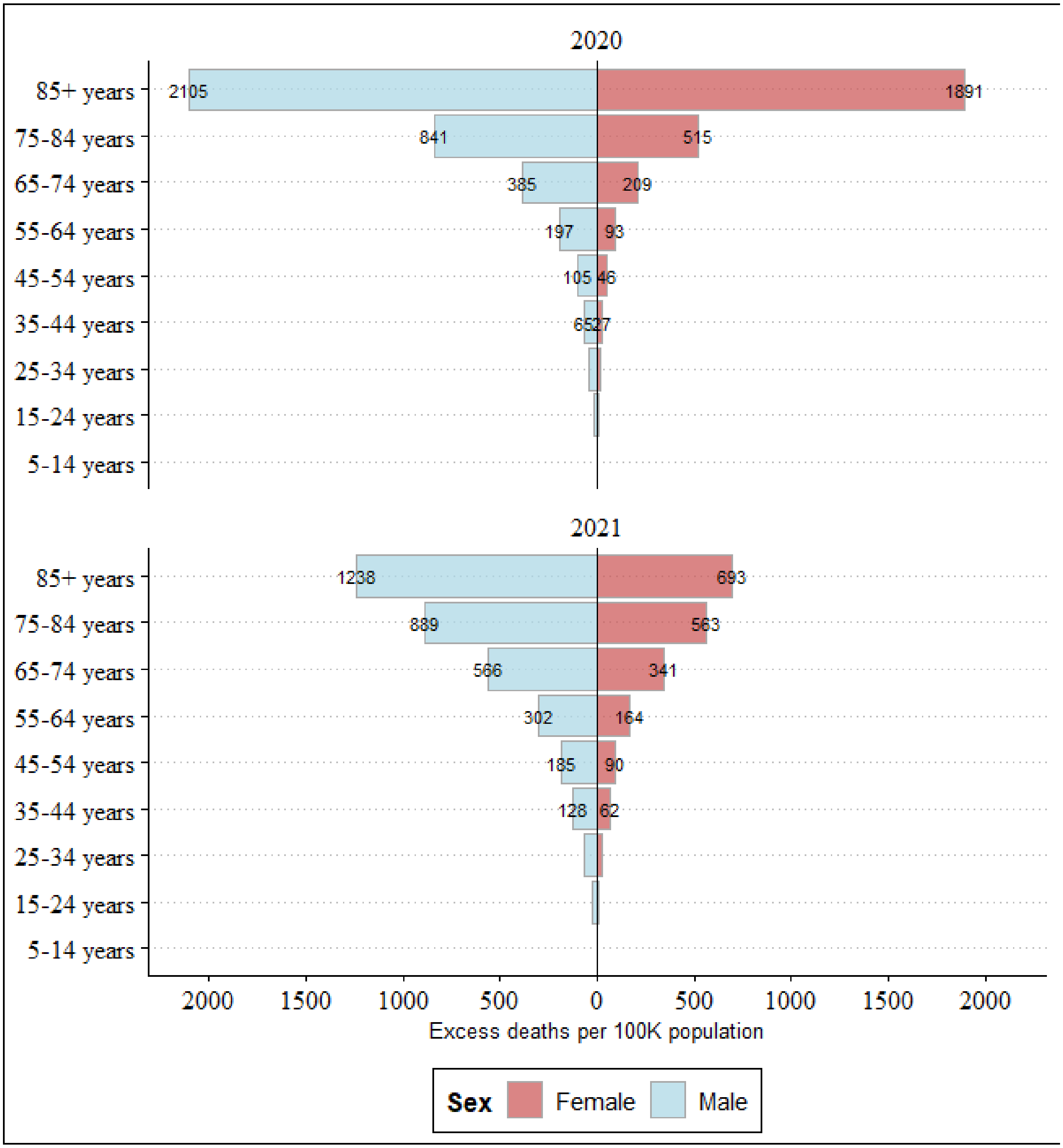


**Figure ED1.** Excess mortality estimates stratified by age and sex, during 2020 (top) and 2021 (bottom).

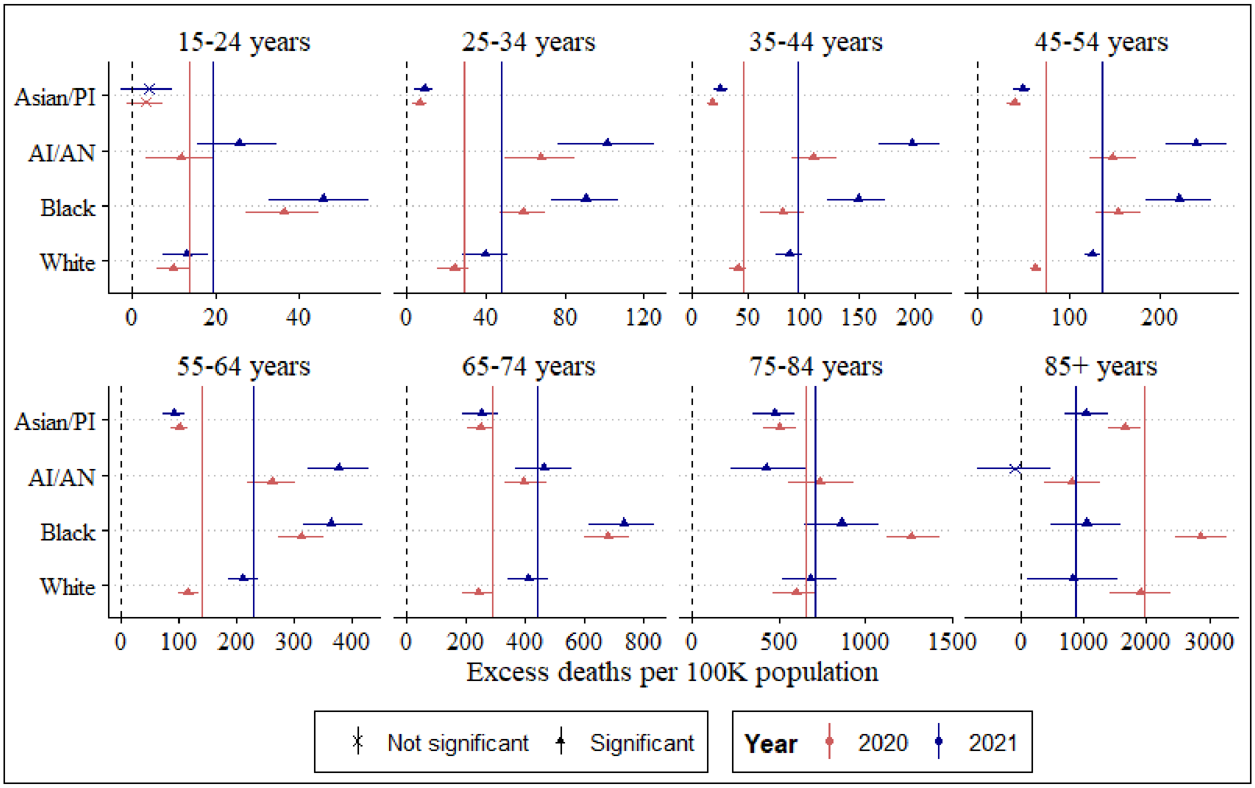


**Figure ED2.** Excess mortality estimates stratified by age and race. The vertical lines denote the age-specific national rate across all races for 2020 (red) and 2021 (blue). Instances where the lower 95% uncertainty bound was negative were not considered to be significant (denoted by a *x*; for example, 15-24 years among Asian/PI). Estimates for none of the race groups among 5-14 year olds were significant, and hence not shown.

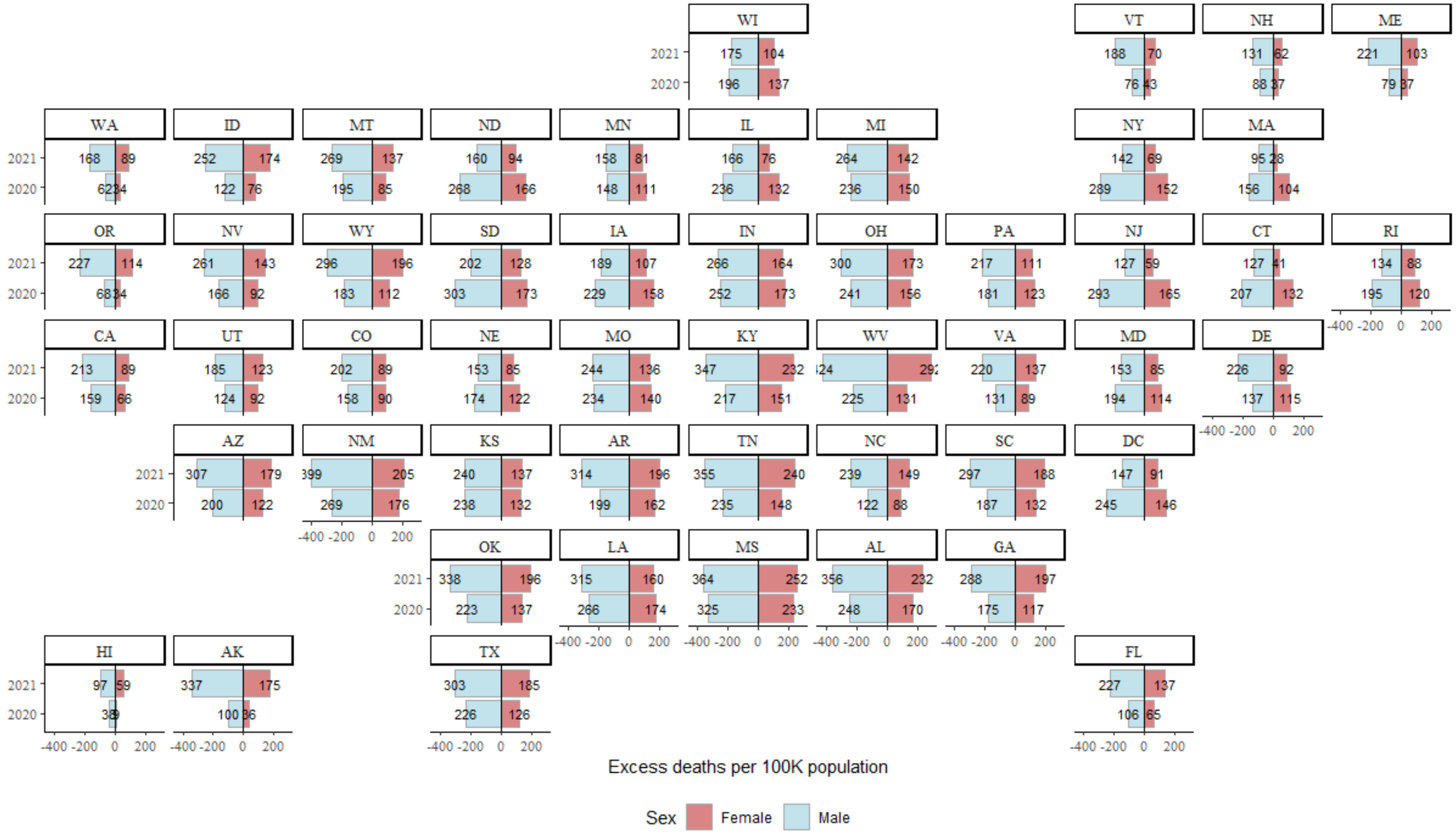


**Figure ED3.** State excess mortality estimates in 2020 and 2021, stratified by sex. Estimates are age-standardized to national age distribution using the direct method.

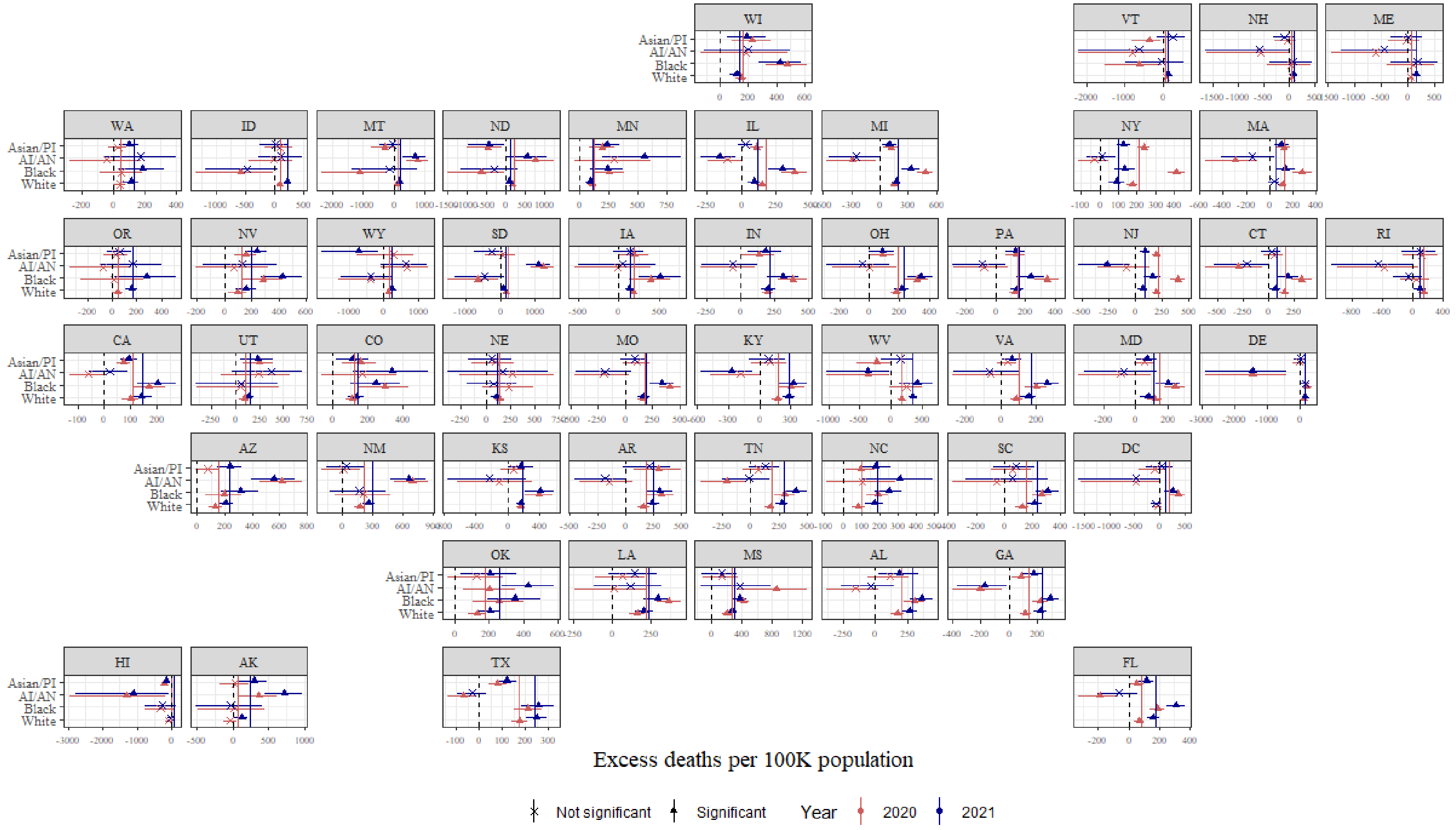


**Figure ED4.** State excess mortality estimates in 2020 and 2021, stratified by race. Estimates are age-standardized to national age distribution using the direct method. The solid vertical lines denote state overall estimates. Instances where the 95% uncertainty bounds have opposite signs were not considered to be significant (denoted by a solid circle; for example, AI/AN in New York).

| | Group quarters, Pr. | Uninsured, Pr. | No vehicle, Pr. | Single parent, Pr. | Age <17, Pr. | Minority, Pr. | Crowding, Pr. | No high school, Pr. | Unemployment, Pr. | Mobile home, Pr. | Limited English prof., Pr. | Poverty, Pr. | Multiple unit bldgs., Pr. | Per capita income | Age >65, Pr. | Bachelors degree, Pr. | Disablility, Pr. |
|---|---|---|---|---|---|---|---|---|---|---|---|---|---|---|---|---|---|
| Pop. density | -0.02 | -0.2 | 0.13 | 0.15 | -0.04 | 0.14 | -0.04 | -0.22 | 0.06 | -0.45 | 0.28 | -0.19 | 0.59 | 0.4 | -0.42 | 0.51 | -0.33 |
| Disablility, Pr. | 0.01 | 0.25 | 0.3 | 0.09 | -0.26 | -0.11 | 0.02 | 0.51 | 0.48 | 0.61 | -0.36 | 0.58 | -0.56 | -0.64 | 0.47 | -0.62 | |
| Bachelors degree, Pr. | 0.05 | -0.37 | -0.18 | -0.18 | -0.07 | 0.04 | -0.18 | -0.7 | -0.34 | -0.66 | 0.25 | -0.53 | 0.7 | 0.77 | -0.26 | | |
| Age >65, Pr. | -0.07 | -0.15 | -0.02 | -0.41 | -0.54 | -0.49 | -0.39 | -0.04 | -0.06 | 0.2 | -0.43 | -0.03 | -0.41 | -0.07 | | | |
| Per capita income | -0.13 | -0.52 | -0.35 | -0.34 | -0.1 | -0.21 | -0.29 | -0.76 | -0.53 | -0.68 | 0.17 | -0.81 | 0.56 | | | | |
| Multiple unit bldgs., Pr. | 0.12 | -0.3 | 0.09 | 0.06 | 0.01 | 0.16 | -0.02 | -0.49 | -0.18 | -0.67 | 0.35 | -0.32 | | | | | |
| Poverty, Pr. | 0.17 | 0.5 | 0.56 | 0.48 | 0.01 | 0.38 | 0.33 | 0.69 | 0.66 | 0.56 | -0.09 | | | | | | |
| Limited English prof., Pr. | 0.01 | 0.26 | -0.04 | 0.19 | 0.3 | 0.55 | 0.47 | 0.14 | -0.01 | -0.21 | | | | | | | |
| Mobile home, Pr. | -0.05 | 0.48 | 0.06 | 0.09 | -0.03 | 0.09 | 0.25 | 0.63 | 0.39 | | | | | | | | |
| Unemployment, Pr. | 0.03 | 0.32 | 0.47 | 0.42 | -0.04 | 0.4 | 0.25 | 0.49 | | | | | | | | | |
| No high school, Pr. | 0 | 0.63 | 0.32 | 0.38 | 0.18 | 0.4 | 0.47 | | | | | | | | | | |
| Crowding, Pr. | -0.05 | 0.54 | 0.09 | 0.35 | 0.42 | 0.53 | | | | | | | | | | | |
| Minority, Pr. | 0.11 | 0.52 | 0.28 | 0.53 | 0.3 | | | | | | | | | | | | |
| Age <17, Pr. | -0.31 | 0.3 | -0.14 | 0.39 | | | | | | | | | | | | | |
| Single parent, Pr. | 0.1 | 0.31 | 0.39 | | | | | | | | | | | | | | |
| No vehicle, Pr. | 0.26 | 0.09 | | | | | | | | | | | | | | | |
| Uninsured, Pr. | -0.06 | | | | | | | | | | | | | | | | |

**Figure ED5.** Pairwise Spearman correlation of SVI variables. A diverging color scheme represents the strength of the correlation from -1 (blue) to 1 (green), with no correlation (0) denoted by white. The lower triangle below the diagonal was suppressed as the correlations are symmetric; the diagonal was also suppressed as correlations equal 1. *Age >65, Pr.*: Prop. of population 65+ years; *Age <17, Pr.*: Prop. of population under 17 years; *Crowding, Pr.*: Prop. of population living in housing units with more people than rooms; *Disability, Pr.*: Prop. of population who have a disability; *Group quarters, Pr.*: Prop. living in group quarters; *Limited English prof., Pr.*: Prop. who speak English "less than well"; M*inority, Pr.*: Prop. who are minorities (all except White, non-Hispanic); *Mobile home, Pr.*: Prop. of housing that are mobile homes; *Multi unit bldgs.., Pr.*: Prop. of housing that is in structures with 10 or more units; No *high school, Pr.*: Prop. without high school diploma; *No vehicle, Pr.*: Prop. who do not have a vehicle; *Per capita income*: Per capita income of the population, in 100,000 US dollars; *Pop density*: Population per sq. mile, log; *Poverty, Pr.:* Prop. below poverty; *Bachelors degree, Pr.*: Prop. with Bachelor's degree or higher; *Single parent, Pr.*: Prop. of households with single parent and children under 18; *Unemployment, Pr.*: Prop. who are unemployed: *Uninsured, Pr.*: Prop. who are uninsured.

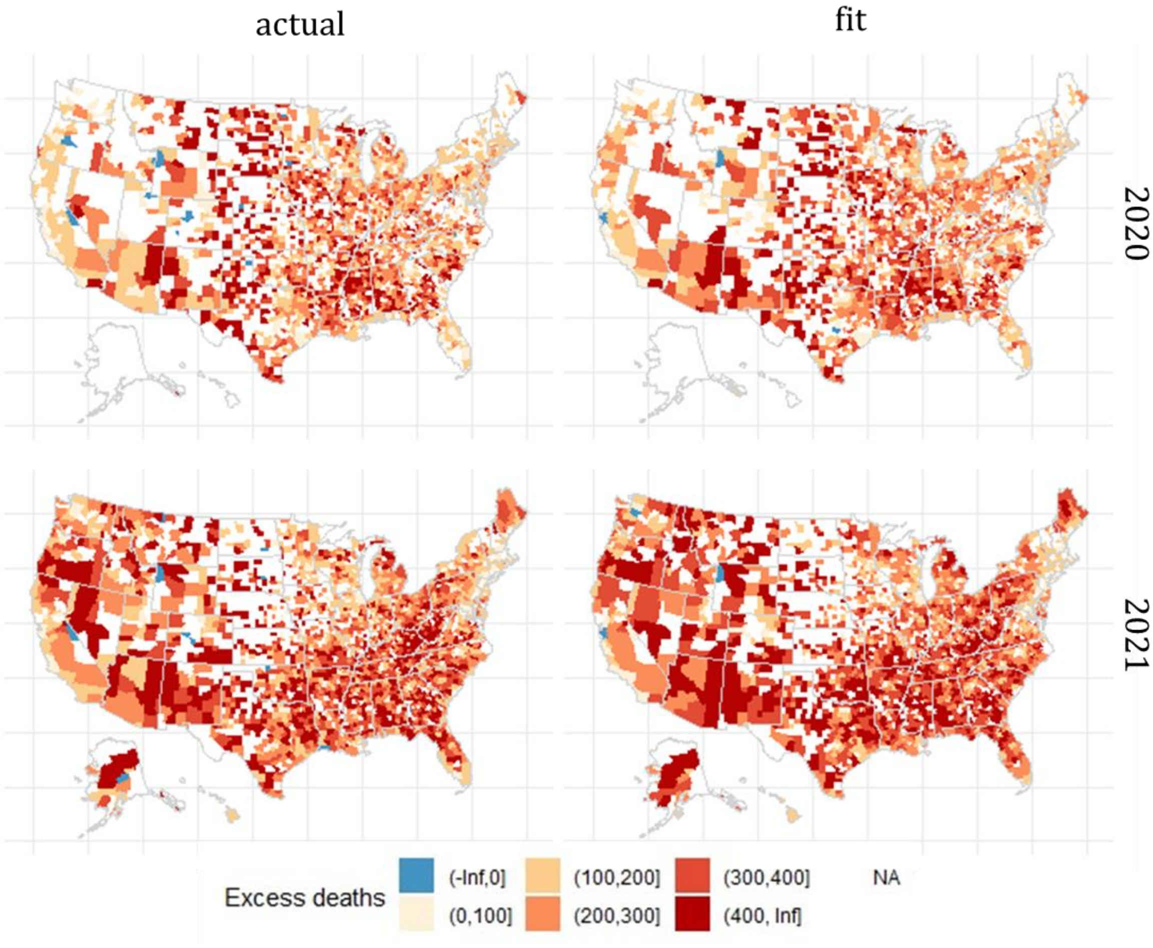


**Figure ED6.** Excess mortality rates used as response in the multivariate model (*actual*) and the model fit (*fit*) during 2020 (top row) and 2021.

*Appendix Text 1: Mismatch in race categories*

The race variable employed in the detailed mortality dataset for the years 2003 to 2020 used four categories: White, Black, American Indian/Alaskan Native (AI/AN) and Asian/Pacific Islander (Asian/PI). For 2021, for which detailed data were unavailable at the time the analysis was undertaken, and provisional data from CDC Wonder Multiple cause of death (MCD) interface were used, the only available race variable used five categories with an additional category called 'more than one race'.

In other words, decedents who would be assigned to one of the four race categories in the detailed mortality dataset were assigned to the 'more than one race' category in the MCD. To assess the impact of this difference in categorization, we examined aggregate counts for the race groups under the two race variables (with 4 categories and 5 categories) during 2020. We found that 7.2% of AI/AN decedents and 4.4% of Asian/PI decedents were classified as 'more than one race' by the MCD. This is higher than the percentages for White (0.3%) and Black (0.6%) decedents.

Hence, the 2021 excess mortality estimates reported in the manuscript for these two race groups are not directly comparable to their corresponding 2020 rates, and caution is warranted in interpreting the change in their excess rates.

| Race group | Detailed Mortality | MCD | Difference (%) |
|---|---:|---:|---:|
| White | 2779151 | 2771478 | 7673 (0.28) |
| Black | 452121 | 449458 | 2663 (0.59) |
| AI/AN | 27976 | 25969 | 2007 (7.17) |
| Asian/PI | 100930 | 96523 | 4407 (4.37) |
| More than one | NA | 17067 | NA |

**Table S1.** Distribution of decedents in 2020 by racial categories in two data sources.

*Appendix text 2. Age standardization*

When comparing mortality rates across time or between groups, it is necessary to ensure that the age composition in the two groups is similar. In the US, there are known differences in age distributions across states (for example, age distribution skews older in Florida compared to most other states), by sex (women live longer than men) and race. As such, age standardization was required in this study to control for differences in age distributions and to render comparisons meaningful. The direct method of age standardization computes the mortality rates that would occur in a comparison group (say a state) if the observed age-specific death rates in the group were present in a population with the same age distribution as a reference population. Here, we used the US national age distribution in 2020 as the reference population.

The direct method has been implemented in the *epitools* package (1) in *R* (2) based on a method described by Anderson and Rosenberg (3). Briefly, using state excess mortality rates as an example, let $p_s^a$ denote the population in age group $a$ and state $s$ and $d_s^a$ the corresponding excess deaths. Age-specific excess mortality rate in state $s$ for age group $a$ is hence $d_s^a/p_s^a$, and the unadjusted mortality rate for state s is given by $\frac{\sum_a d_s^a}{\sum_a p_s^a}$. The state's age-standardized mortality rate is given by $\sum_a \left(\frac{d_s^a}{p_s^a} * \frac{\sum_s p_s^a}{\sum_{a,s} p_s^a}\right)$, which can be seen as a weighted average where the weight, $\frac{\sum_s p_s^a}{\sum_{a,s} p_s^a}$, is the proportion of the national population ($\sum_{a,s} p_s^a$) that belongs to age group $a$ ($\sum_s p_s^a$).

Age-standardized rates among men and women and for the four race groups nationally were calculated analogously. Figure S1 shows the change in the excess mortality rate due to age-standardization. When standardized, excess mortality rates decreased among women and increased among men, as expected. The magnitude of change was larger in Black and AI/AN groups. As a result, the difference between White and Black groups increased considerably with standardization; and, while the crude rates of White and AI/AN were similar, standardized rates among AI/AN were larger than for the White population. We were not able to report standardized estimates at county-level because disaggregating mortality counts by age at the county-level would have led to significant data suppression in the provisional dataset for 2021.

*Appendix text 3: Temporal cross validation*

To inspect model estimates of expected deaths during the years before the pandemic, we performed a temporal cross-validation exercise. For each of the years 2010-2019, we trained the models (as described in the main text) with data up to (and not including) the candidate year and projected deaths one year ahead. As the response being modeled is all-cause deaths, a mismatch between expected and observed deaths does not necessarily indicate sub-optimal quality of the models, but rather possible excess deaths from other causes (for example, drug overdose deaths). Nevertheless, inspecting the concurrence between the observed and predicted deaths is helpful to check for consistent biases in model estimates.

Figure S2 shows uncertainty bounds of the expected all-cause mortality rates against observed rates for the US overall, and disaggregated by age, race and sex. National expected death estimates were within the 95% uncertainty bounds in 7 of the 10 years, and were overestimates in 2 years (2014 and 2016) and underestimates in 1 year (2019). Similar pattern was observed in estimates for men and women and in 3 of the 4 race groups. Estimates for AI/AN were biased higher in a majority of the years, suggesting that one or more of the 18 models for this race group was unable to capture the temporal or spatial trends. While reasons for underperformance were not thoroughly investigated, one potential cause could be the relatively sparse distribution of AI/AN across counties.

Similar higher bias was observed in estimates for the 45-54 year age group, but encouragingly, expected death estimates for the 3 older age groups with the highest mortality rates (65-74 years, 75-84 years and 85+ years) appear to capture the clear trend in mortality over the training period.

At the county-level we calculated the coverage of the estimates in each year i.e. the proportion of the counties whose observed rates were within the 95% uncertainty interval of the model estimated expected rates (Figure S3). A coverage close to .95 is optimal. Results show good coverage at the start of the validation period (2010-12) with degradation over time. Across all years the coverage was 89%. A similar trend in coverage was observed with interval bounds of 90% and 50%.

*Appendix text 4*: *Comparison with NCHS excess mortality estimates*

We compared national and state excess mortality estimates from the method described here with estimates published by the National Center for Health Statistics (NCHS) (4). National NCHS' estimates, at 133 and 162 excess deaths per 100,000 population in 2020 and 2021, respectively, were lower than our estimates  (149 and 175, respectively). At the state level, strong correlation between the two estimates was observed (Spearman's *rho*: 0.76 in 2020; 0.93 in 2021). NCHS' estimates were lower than our estimates in 36 states during 2020 and 45 states during 2021 (Figure S4). In 91% of instances (93 of 102 state and year combinations), NCHS estimates were within the 95% uncertainty bounds estimated by our method.

*Appendix Text 5: Associations between excess mortality and socioeconomic vulnerability*

Results were presented in the main text for associations when limiting to those counties with statistically significant estimates. Here we report effect sizes when this condition was relaxed and all counties were included.

Linear univariate models, adjusting for deaths from Covid-19, showed strong associations of all-cause excess mortality rates with most of the social vulnerability measures considered. Prevalence of single parent households with children (effect estimate per standard deviation: mean=29.6; 95% CI: 24-35), unemployment (29.1; 24-34), poverty (27.2; 22-33), disability (26.6; 21-32), and not having access to a personal vehicle (19.1; 14-25) were estimated to have the highest positive associations, each measure explaining about 40% of the variability in excess deaths during 2020. Per capita income (-30.4; -36--25) and higher education (Bachelor's degree or above; -27.7; -33--22) were found to be strongly protective. The magnitude of effect was higher in 2021 than in 2020 for nearly all measures, although a decrease in $R^2$ was observed.

Linear multivariate models with all measures included together as explanatory variables explained a slightly higher (45%) variability in both years. Of interest is an increase in the effect estimate of population density relative to the univariate model during both years.

*Appendix Text 6: Associations between Covid-19 mortality and socioeconomic vulnerability*

Results were presented in the main text for associations between all-cause excess mortality and socioeconomic vulnerability. Here we report effect sizes with deaths certified to be from Covid-19 (ICD-10 underlying cause of death code: U07.1).

Linear univariate models found strong associations with Covid-19 mortality rates for most of the social vulnerability measures considered (Figure S5). To limit to the three measures with the highest positive and the highest negative associations in 2020, prevalence of those who have not completed high school (effect estimate per standard deviation: mean=26.6; 95% CI: 23-30), of those who are uninsured (21.6; 18-25), and those living in poverty (21; 18-24) were estimated to have the highest positive associations, whereas higher education (Bachelor's degree or above; -31.1: -34 - -28 ), population density (-28.3; -31 - -25), and per capita income (-24.8; -28 - -22) and were found to be strongly protective. For a majority of the variables, the magnitude of effect was higher in 2021 than in 2020.

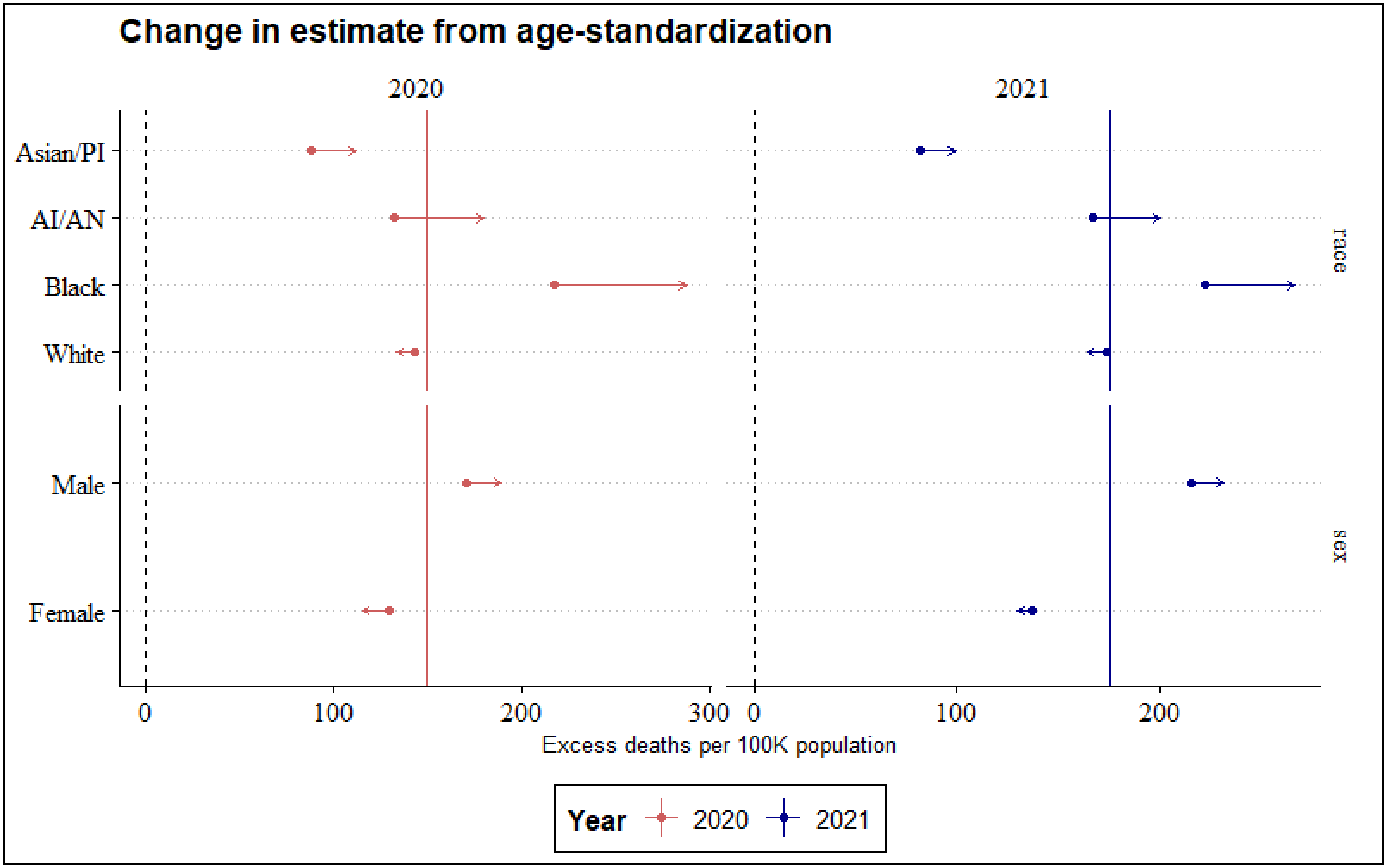


**Figure S1.** Change in national excess mortality for 4 racial groups, men and women due to age-standardization with the direct method using overall national population distribution as reference. Data points denote unadjusted rates and arrow ends denote the standardized rate (as reported in Figure 1b).

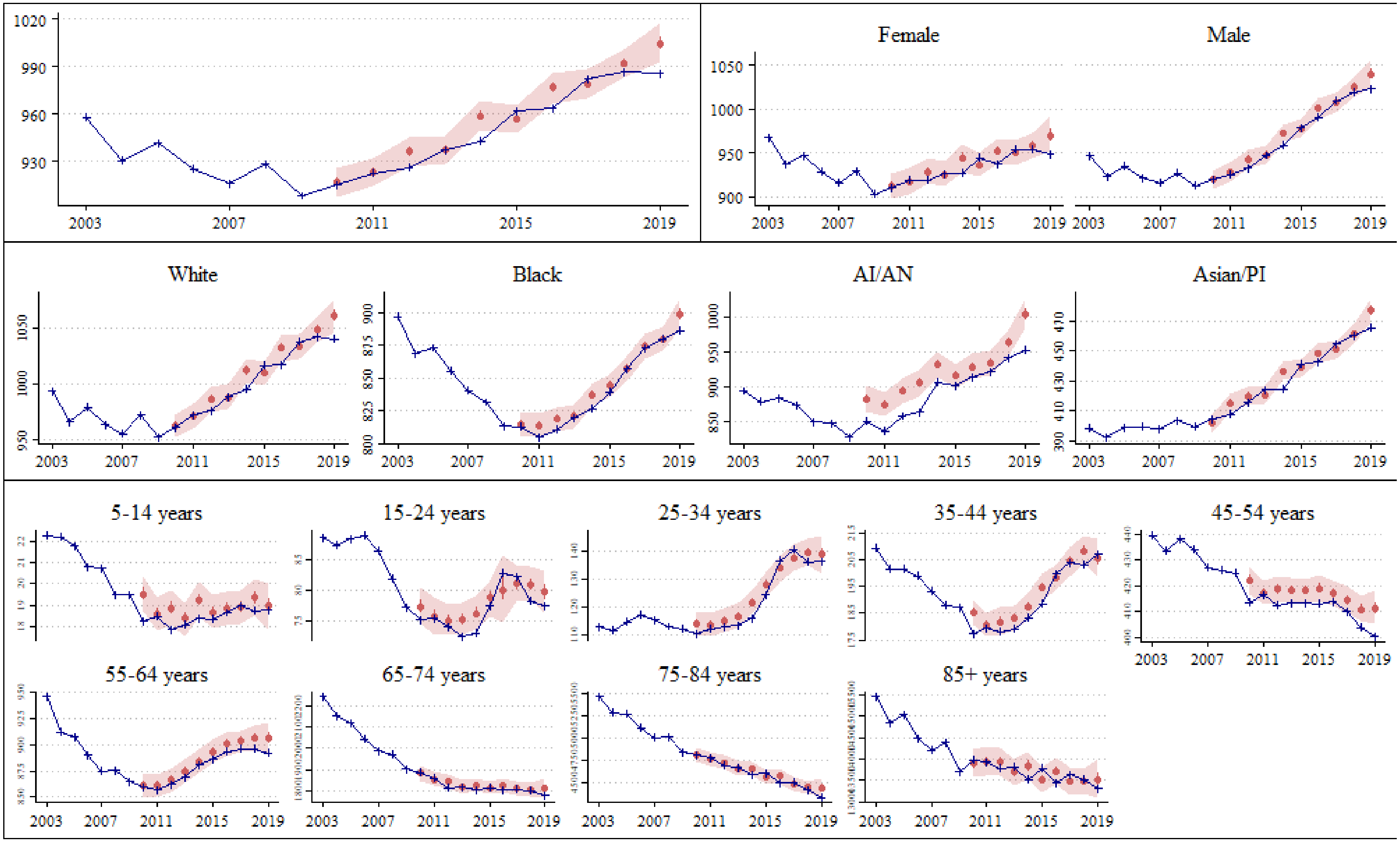


**Figure S2.** Cross validation for pre-Covid period 2010 – 2019, US national (top row, left), stratified by sex (top row, right), race (middle row) and age (bottom two rows). Observed deaths (in blue) and predicted median (red point), and 95% uncertainty interval (red shaded region) of expected deaths per 100,000 population.

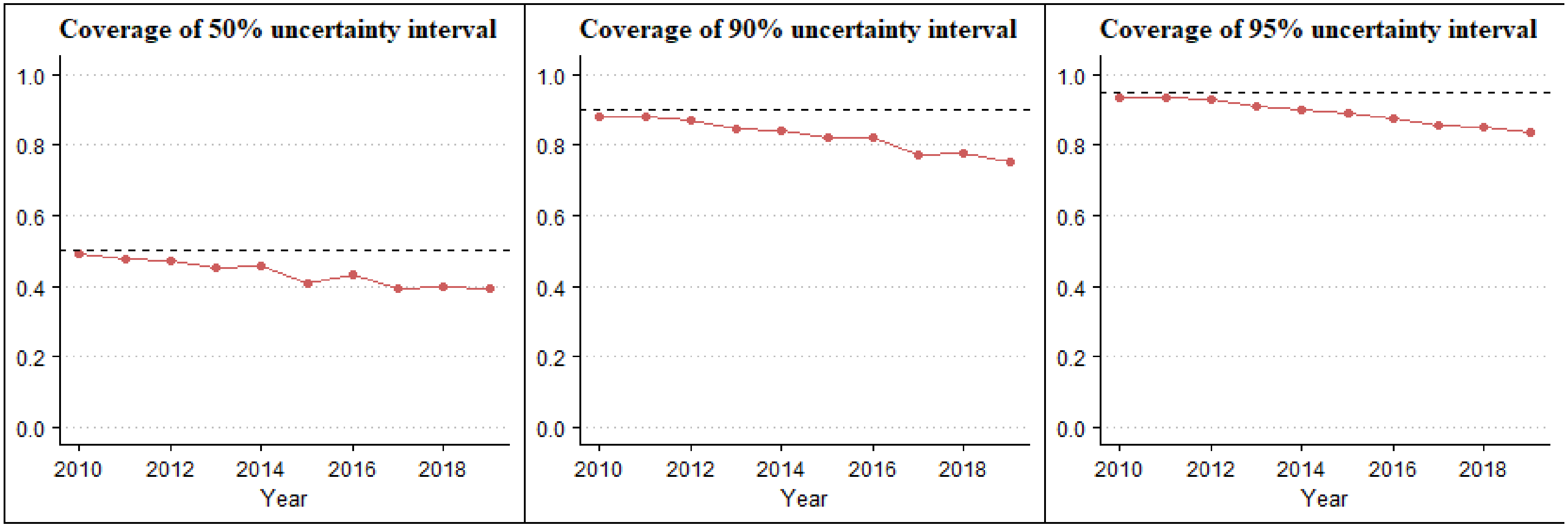


**Figure S3.** Coverage of model uncertainty interval in temporal cross validation at county-level.

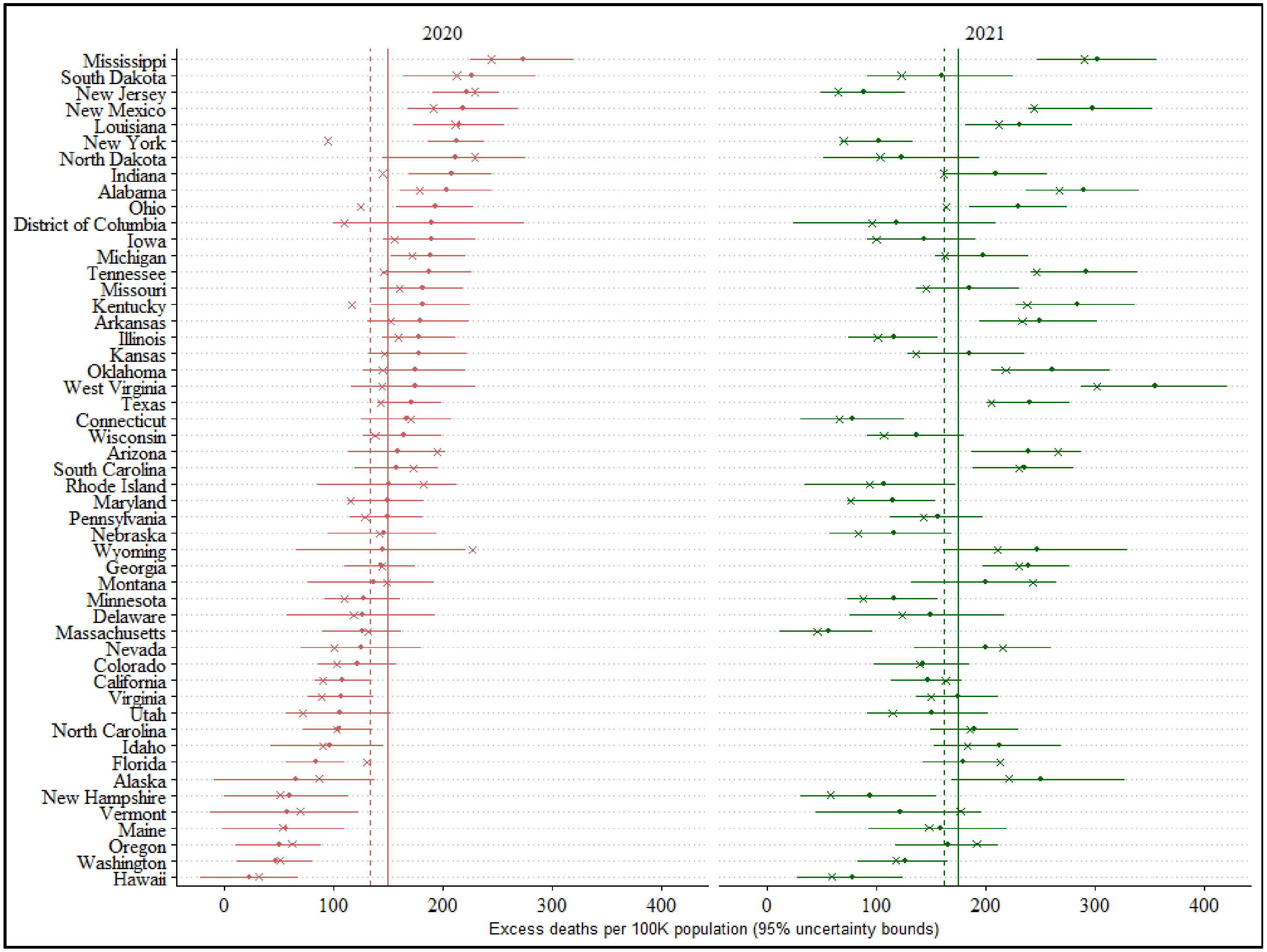


**Figure S4.** Comparison of excess mortality estimates from NCHS and the current study. NCHS estimates are denoted by 'x' and our estimates are denoted by the point (median) and bars (95% uncertainty interval). Vertical lines denotes national excess estimate from this study (solid) and NCHS (dashed).

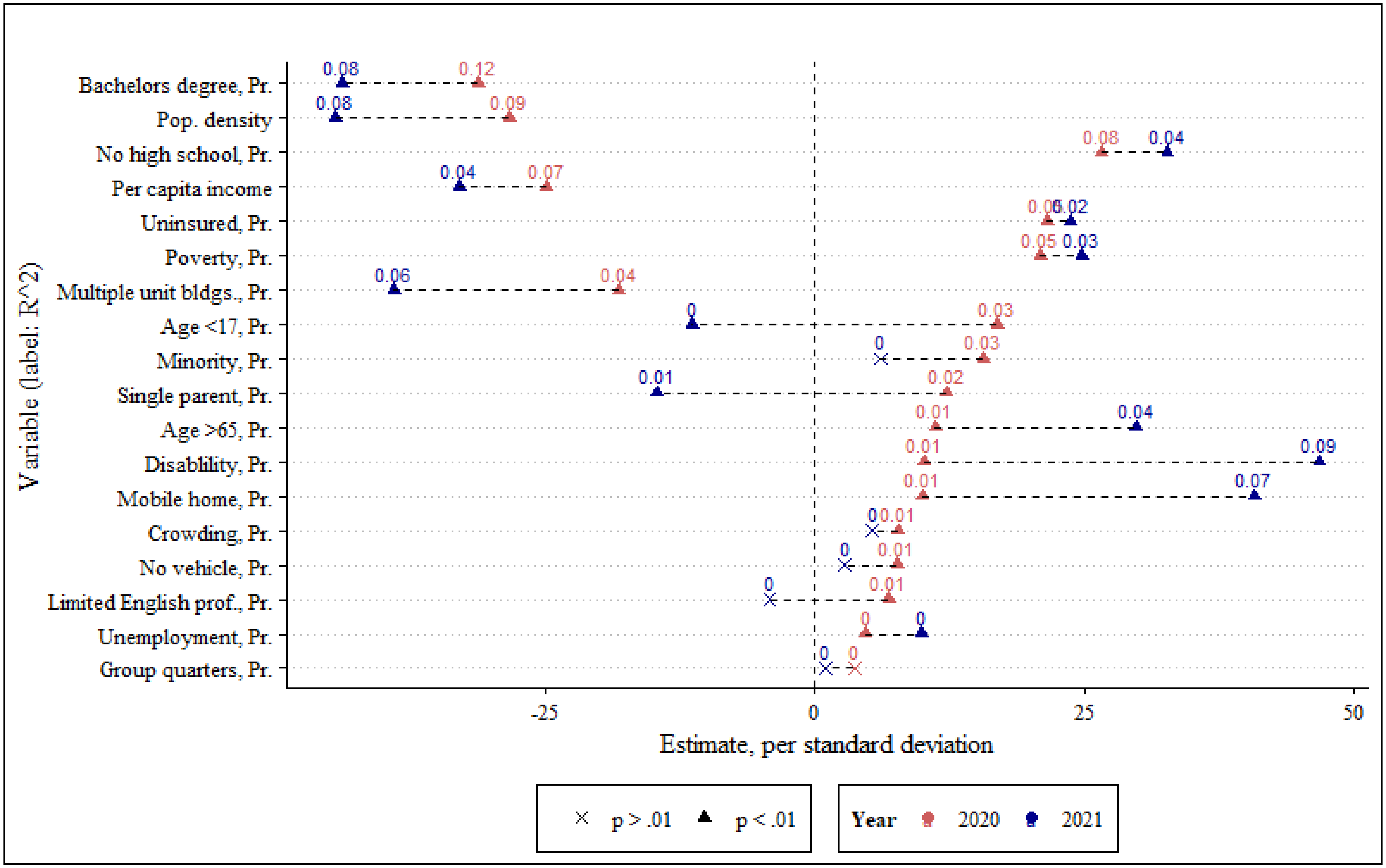


**Figure S5.** Standardized effect estimates (per standard deviation) from linear univariate models of social vulnerability measures and Covid-19 mortality during 2020 (red) and 2021 (blue). Non-significant effect estimates are denoted by *x* ($p > .01$). The data points are labeled with estimated $R^2$, and the measures are ordered by absolute effect in 2020 to least. See Table 1 for measure descriptions. Proportions are abbreviated as *Pr.*
